# A clinical trial to study changes in neural activity and motor recovery following brain-machine interface enabled robot-assisted stroke rehabilitation

**DOI:** 10.1101/2020.04.26.20077529

**Authors:** Nikunj Bhagat, Nuray Yozbatiran, Jennifer L. Sullivan, Ruta Paranjape, Colin Losey, Zachary Hernandez, Zafer Keser, Robert Grossman, Gerard Francisco, Marcia K. O’Malley, Jose Contreras-Vidal

**Affiliations:** Non-Invasive Brain Machine Interface Systems Laboratory, University of Houston; Department of Physical Medicine and Rehabilitation, McGovern Medical School, NeuroRecovery Research Center at TIRR Memorial Hermann, University of Texas Health Science Center at Houston; Mechatronics and Haptic Interfaces Laboratory, Rice University; Houston Methodist Research Institute; NSF IUCRC BRAIN, University of Houston

## Abstract

**Background:** Brain-machine interfaces (BMI) based on scalp electroencephalography (EEG) have the potential to promote cortical plasticity following stroke, which has been shown to improve motor recovery outcomes. However, clinical efficacy of BMI-enabled robotic rehabilitation in chronic stroke population is confounded by the spectrum of motor impairments caused by stroke.

**Objective:** To evaluate the efficacy of neurorehabilitation therapy on upper-limb motor recovery, by quantifying changes in clinical, BMI-based, and kinematics-based metrics. Further, to identify neural correlates or biomarkers that can predict the extent of motor recovery.

**Methods:** Chronic stroke survivors (n = 10, age 55 ± 9.2y, chronicity 3.1 ± 2.8y) were recruited to participate in a 4-6 weeks long clinical study. Participants completed 12 therapy sessions that involved a BMI enabled powered exoskeleton (MAHI Exo-II) for training, which targeted elbow flexion and extension. Clinical assessments including Fugl-Meyer Upper Extremity (FMA-UE) and Action Research Arm Test (ARAT) were measured up to 2-months after therapy. BMI performance, kinematic performance, and change in movement related cortical potentials (MRCP) were also determined.

**Results:** On average, 132 ± 22 repetitions were performed per participant, per session. BMI accuracy across all sessions and subjects was 79 ± 18%, with a small number of false positives (23 ± 20%). FMA-UE and ARAT scores improved significantly over baseline after therapy and were retained at follow-ups (ΔFMA-UE = 3.92 ± 3.73 and ΔARAT = 5.35 ± 4.62, *p* < 0.05). 80% participants (7 with moderate-mild impairment and 1 with severe-moderate impairment) reached minimal clinically important difference (MCID: FMA-UE > 5.2 or ARAT > 5.7) during the course of the study. Kinematic measures indicate that, on average, participants’ movements became faster and smoother. Quantification of changes in MRCP amplitude showed significant correlation with ARAT scores (ρ = 0.72, *p* < 0.05) and marginally significant correlation with FMA-UE (ρ = 0.63, *p* = 0.051), suggesting higher activation of ipsi-lesional hemisphere post-intervention. The study did not have any adverse events.

**Conclusion:** This study presents evidence that BMI enabled robotic rehabilitation can promote motor recovery in individuals with chronic stroke, several years after injury and irrespective of their impairment level, or location of the lesion (cortical/subcortical) at baseline. Further, the extent of motor recovery was correlated with changes in movement related potentials, occurring contralateral to the impaired arm.

**Support:** NIH National Robotics Initiative Grant R01NS081854 and a grant from Mission Connect, a project of TIRR Foundation.

## 1. Introduction

Upper-limb motor weakness occurs in 77% of first time and 55 – 75% chronic stroke survivors and significantly affects their quality of life (Coscia et al., 2019; Lawrence et al., 2001). Regaining arm and hand function is an essential part of achieving independence in daily life and therefore is a major goal of rehabilitation programs. While most traditional rehabilitative strategies are using bottom-up approaches by incorporating training of distal body parts to influence neural systems (Belda-Lois et al., 2011), e.g., constraint induced movement therapy (CIMT) (Wolf et al., 2008), robotic arm training (Lo et al., 2010), bilateral arm training (Whitall et al., 2000), or functional electrical stimulation (Makowski et al., 2014), a number of studies have addressed clinical effects of top-down approaches, e.g., brain stimulation (Dimyan & Cohen, 2010; Liew et al., 2014), motor imagery (Lopez et al., 2019) and brain-computer interface (BCI) (Daly & Wolpaw, 2008) to induce neuroplastic changes in the sensorimotor network, especially in stroke survivors with severe motor deficits.

Brain-machine/computer interface (BMI/BCI) can improve treatment benefits when combined with robotic and muscular stimulation based neurorehabilitation therapies, by capitalizing on the principles of Hebbian plasticity (Soekadar et al., 2015). Indeed, previous clinical studies that combined motor imagery based BMIs with upper-limb arm and hand exoskeletons or electrical muscle stimulation achieved significantly better motor improvement compared to sham or control groups (Ang et al., 2014; Biasiucci et al., 2018; Frolov et al., 2017; Pichiorri et al., 2015; Ramos-murguialday et al., 2013).

Despite these promising findings, evidence of cortical changes following neurorehabilitation therapy remain largely unproven, and a neural correlate (or biomarker) that can predict the extent of motor recovery still remains elusive. To address this deficit, (Ramos-murguialday et al., 2013) used functional MRI and found post-therapy activations in the ipsi-lesional motor and pre-motor cortices to be correlated (ρ = 0.55) with Fugl-Meyer Assessment for Upper Extremity (FMA-UE) scale. (Ang et al., 2014) found the revised Brain Symmetry Index to be inversely correlated to motor improvement, suggesting that bilateral activations of cortical hemispheres led to better recovery (ρ = -0.62). Others have reported increased resting state functional connectivity and integrity of white matter tracts (via diffusion tensor imaging) within the motor areas of both hemispheres following BMI mediated stroke rehabilitation (Biasiucci et al., 2018; Rathee et al., 2019; Song et al., 2015).

In this study, we explored the relationship between movement related cortical potentials (MRCPs) and motor recovery following 12 sessions of BMI-enabled robot-assisted stroke rehabilitation. It was hypothesized that MRCP amplitude and latency (i.e., duration of MRCP prior to movement onset) would increase, on account of increased activation of the ipsi-lesional hemisphere or inhibition of competing contra-lesional hemisphere, following motor relearning and cortical reorganization (Yilmaz et al., 2015). Further, to increase patient engagement and strengthen MRCPs, the BMI algorithms were optimized to detect MRCPs in single-trials using our previously published method (Bhagat et al., 2016). Preliminary findings of our clinical trial, reporting the improvements in movement quality and arm function from initial 6 participants, were published in (J. L. Sullivan et al., 2017). In this paper, we present a comprehensive analysis from 10 participants by determining longitudinal efficacy of EEG-based BMIs, as well as by evaluating changes in brain activity, motor recovery, and movement quality following BMI-exoskeleton therapy.

## 2. Methods

A single-arm clinical study (ClinicalTrials.gov #NCT01948739) was conducted to evaluate the efficacy of BMI enabled exoskeletons on stroke recovery and brain activity. The study procedures were approved by the Institutional Review Boards of University of Houston, Rice University, University of Texas Health Science Center at Houston, and the Houston Methodist Hospital at Houston, Texas. All participants provided informed consent in accordance with the Declaration of Helsinki.

### A Study Participants

Between 2013 and 2018, 160 individuals were screened for eligibility based on following inclusion criteria: first time subacute and chronic stroke (i.e. at least 3 months since injury); stable baseline arm function (see below); hemiparesis of upper extremity (manual muscle testing of at least 2 but no more than 4 out of 5 in elbow and wrist flexors); no joint contracture or severe spasticity; no neglect that would preclude participation in the training protocol; presence of proprioception; no history of neurolytic procedure in the past four months; and no contraindication to MRI. Persons with orthopedic limitation of upper extremity that would affect motor performance; lack of motivation due to untreated depression were excluded from the study. To evaluate baseline arm function stability, FMA-UE assessment was performed at screening and was repeated one month later. A participant was enrolled only if the difference in FMA-UE scores at these visits was ≤ 3 points (Klamroth-Marganska et al., 2014).

Among the participants excluded at screening (n = 142), 117 did not meet the inclusion criteria, 4 did not have a stable baseline, and 13 declined to participate. In addition, 8 individuals that previously participated in our pilot study (Bhagat et al., 2016) for the clinical trial were excluded, since they were familiar with BMI-exoskeleton therapy paradigm. Subsequently, eighteen participants enrolled in the study and were assigned to the BMI-exoskeleton therapy group, and there was no control group. Among these participants, 10 individuals completed the protocol. Participants who dropped out of the study had MRI contraindication (n=4), could not commit time to participate in all therapy and assessment sessions (n=3), or were not interested in participating (n=1). The enrollment and intervention details following the Consolidated Standards of Reporting Trials (CONSORT) flow diagram are shown in Fig. 1.

**Fig. 1.**
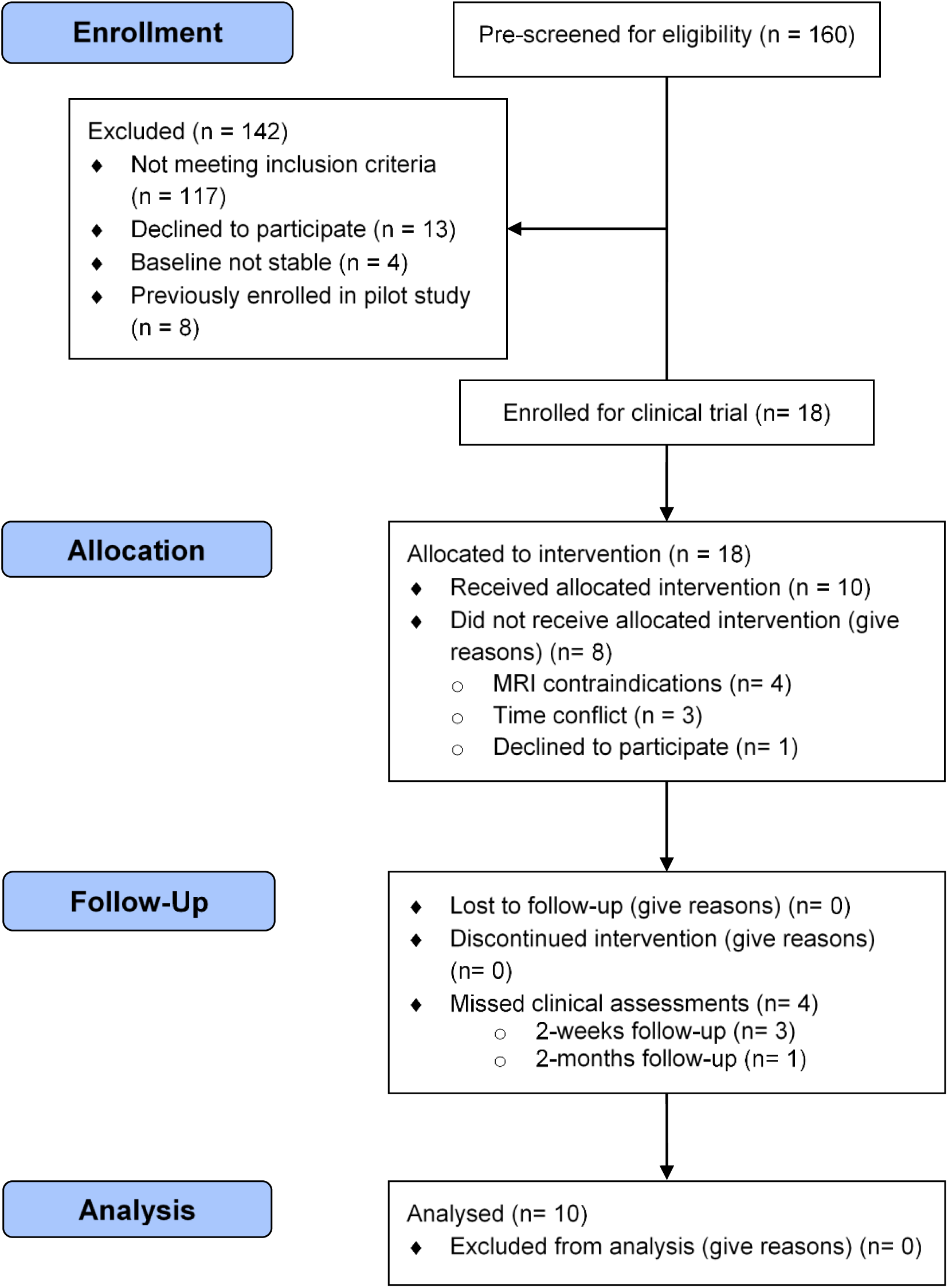
CONSORT flow diagram showcasing patient recruitment, intervention and follow-ups

The study cohort consisted of participants with either cortical (n = 4), subcortical (n = 4), or both cortical and subcortical lesions (n = 2). Specific details regarding the location of stroke lesions, as determined by physicians after reviewing T1-weighted MRI scans, are provided in supplementary materials (Table S1). And in Table 1 below, we present demographics and baseline characteristics of participants who completed the study. Additionally, the average grip and pinch strengths for our participant pool were 11.13 ± 8.7kg and 4.48 ± 2.3kg, respectively. According to (Woytowicz et al., 2017) classification of impairment severity, the participants can be further grouped as severe-moderate impaired (baseline FMA-UE ∈ [16, 34]) or moderate-mild impaired (baseline FMA-UE ∈ [35, 53]), which is also highlighted in Table 1.

**Table 1.**
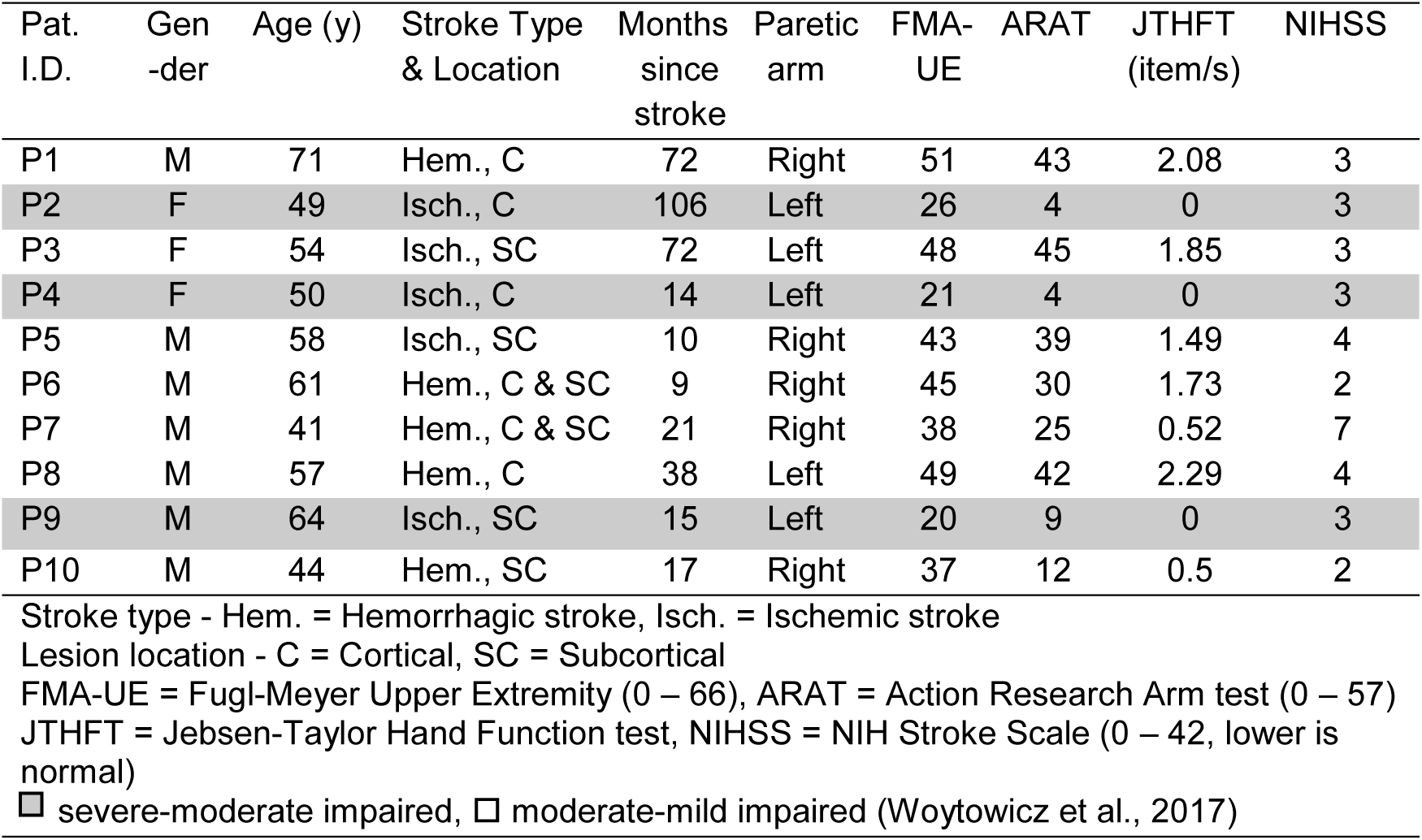
Demographics and baseline assessments of study participants

| Pat.<br>I.D. | Gen<br>-der | Age (y) | Stroke Type<br>& Location | Months<br>since<br>stroke | Paretic<br>arm | FMA-UE | ARAT | JTHFT<br>(item/s) | NIHSS |
| --- | --- | --- | --- | --- | --- | --- | --- | --- | --- |
| P1 | M | 71 | Hem., C | 72 | Right | 51 | 43 | 2.08 | 3 |
| P2 | F | 49 | Isch., C | 106 | Left | 26 | 4 | 0 | 3 |
| P3 | F | 54 | Isch., SC | 72 | Left | 48 | 45 | 1.85 | 3 |
| P4 | F | 50 | Isch., C | 14 | Left | 21 | 4 | 0 | 3 |
| P5 | M | 58 | Isch., SC | 10 | Right | 43 | 39 | 1.49 | 4 |
| P6 | M | 61 | Hem., C & SC | 9 | Right | 45 | 30 | 1.73 | 2 |
| P7 | M | 41 | Hem., C & SC | 21 | Right | 38 | 25 | 0.52 | 7 |
| P8 | M | 57 | Hem., C | 38 | Left | 49 | 42 | 2.29 | 4 |
| P9 | M | 64 | Isch., SC | 15 | Left | 20 | 9 | 0 | 3 |
| P10 | M | 44 | Hem., SC | 17 | Right | 37 | 12 | 0.5 | 2 |

Stroke type - Hem. = Hemorrhagic stroke, Isch. = Ischemic stroke Lesion location - C = Cortical, SC = Subcortical FMA-UE = Fugl-Meyer Upper Extremity (0 – 66), ARAT = Action Research Arm test (0 – 57) JTHFT = Jebsen-Taylor Hand Function test, NIHSS = NIH Stroke Scale (0 – 42, lower is normal) ■ severe-moderate impaired, □ moderate-mild impaired (Woytowicz et al., 2017)

### B Study Protocol and Experiment Design

The clinical trial protocol consisted of 14-15 sessions and 5 functional assessments (Fig. 2A). The initial 2 sessions were used for calibrating the BMI algorithm (see Sec. 2C) to each participant. Participants P2 and P8 underwent an additional calibration session to fine-tune the BMI classifier’s parameters. Once calibrated, the BMI-exoskeleton therapy was provided for 12 sessions, 3 times per week, for 4 weeks. Participant P10 was unavailable during weekdays and hence, his sessions were conducted on the weekends for 6 weeks. The functional assessments were performed twice at baseline as described earlier and once post-treatment, as well as at 2-weeks and 2-months follow-ups. The primary outcome measures were functional improvement in arm and hand movements using FMA-UE test, changes in neural activity as measured by EEG, and improvement in movement quality as determined from the exoskeleton’s kinematics. The secondary outcomes assessed motor recovery using additional clinical scales such as Action Research Arm test (ARAT), Jebsen-Taylor Hand Function test (JTHFT), pinch and grip strengths. FMA-UE score is comprised of 8 scoring items, namely arm movements involving flexor synergy, extensor synergy, combined synergies (e.g. move hand to lumbar spine), out of synergy (e.g. shoulder abduction to 90°, while elbow is at 0° and forearm is pronated), hand, wrist, speed/co-ordination, and reflexes (K. J. Sullivan et al., 2011). Likewise, ARAT scores are the aggregate of 4 subscales: grasp, grip, pinch, and gross movements (Yozbatiran et al., 2008). Additionally, we recorded surface electromyography (EMG) from biceps and triceps muscles of both impaired and unimpaired arms to determine if participants exhibited global synkinesis or motor irradiation (Hwang et al., 2005), but also to provide a ‘ground truth’ for the BMI output (Fig. 2B).

**Fig. 2.**
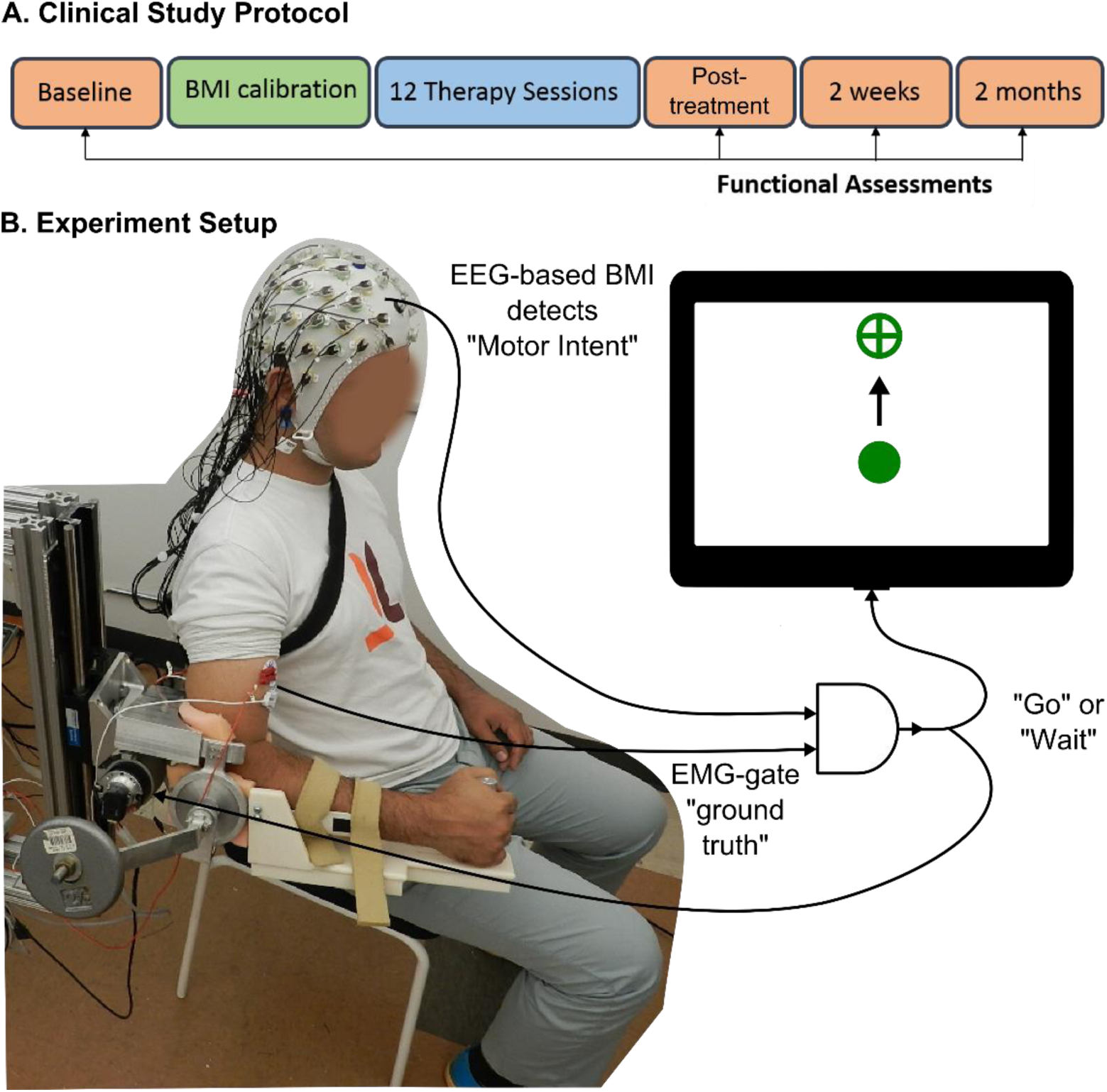
EEG-based BMI control of MAHI exoskeleton for stroke rehabilitation. A) Timeline for the clinical study protocol. B) Schematic representation of the experiment setup, showing a stroke participant’s impaired elbow being trained by the MAHI Exo-II, while EEG and EMG activity are recorded. In this BMI scheme, successful detection of motor intent from EEG is validated against residual EMG activity from impaired arm, before a Go or Wait command is issued to the exoskeleton. A computer screen in front of the participant, cues start and end of trial and provides simultaneous visual feedback of the movement.

Each therapy session lasted 3 to 3.5 hours and included EEG preparation (~45 min.), daily kinematic assessment (~15 min.), therapy time (~ 2 hours), and breaks as needed. During therapy, participants were presented with a center-out reaching task on a computer screen to train their elbow flexion and extension movements, while their impaired arm was supported by the MAHI Exo-II exoskeleton (Fitle et al., 2015). To perform the movement, the participants were instructed to “first think about the movement and then gently attempt to move their arm”. Each trial lasted up to 15 seconds, and the participants could attempt to move multiple times in a trial. If the BMI algorithm successfully detected the motor intention, which was corroborated by EMG activity in the prime muscles, then the exoskeleton was triggered to assist in the movement; otherwise the exoskeleton remained stationary and resisted the movement. This protocol enforced the participants to remain mentally engaged in the task in order to maximize the benefits of the BMI-exoskeleton therapy. Once the target was hit, the exoskeleton automatically returned to center, and after a randomized resting interval (4 – 6s), the next trial was presented. Typically, participants practiced 60 – 180 trials per session, and the number of repetitions increased once they became proficient in controlling the BMI and their fatigue diminished.

### C BMI Algorithm

Our BMI algorithm was based on methods developed previously, wherein an EEG-based classifier’s predictions were gated with residual EMG activity from the impaired arm, before triggering an exoskeleton’s movement (Bhagat et al., 2016). To detect motor intent we identified movement related cortical potentials (MRCPs) from delta-band EEG rhythms (0.1 – 1Hz), using a Go vs. No-go Support Vector Machine (SVM) classifier (Lotte et al., 2007). The classifier was trained for each participant using pre-recorded calibration data, in which they voluntarily moved or triggered movement of the exoskeleton with their impaired arm, while performing motor imagery. Unlike the previous study, wherein we handpicked the EEG channels that were fed to the classifier, here we automated the channel selection process. First, we visually short-listed EEG channels that contained MRCPs from grand averaged movement epochs. Next, we used backward elimination and dropped channels that were less relevant for classification, as determined from the mutual information between class labels and feature vectors (Lan et al., 2005; Peng et al., 2005). The training algorithm also automatically selected the optimal feature extraction window length using ROC curves (Fawcett, 2006). This was achieved by training the classifier offline for different window lengths ranging from 100ms to 1s, in 100ms increments. In each iteration, an ROC curve was obtained using confusion matrices and eventually, the window length corresponding to the classifier with maximum area under ROC curve was considered optimal for that participant.

The online BMI performance was further improved by tuning 2 parameters: the classifier’s prediction probability estimate (*τ_c_*) and number of consecutive Go predictions required before intent is asserted (*N_c_*) (Bhagat et al., 2016). Parameters *τ_c_* and *N_c_* were initially set at 0.5 and 3 respectively, and increased up to 1 and 10 until the participants achieved high accuracy. Once tuned, the BMI classifier and its parameters were fixed for 12 therapy sessions. For configuring the EMG-gate, a simple threshold detection technique was employed. Under this technique, RMS values for EMG signals from impaired hand were baseline corrected by subtracting the mean value over a 30 seconds resting period. The resulting signals were then compared against an empirically determined threshold, typically 5 – 30 units above baseline. The EMG thresholds however, did require to be readjusted between sessions and sometimes within a session, to overcome offsets from poor contact with the skin or from brushing against the exoskeleton’s braces.

### D Computation of Post-treatment MRCP Changes

To quantify changes in neural activity as a result of therapy, we looked at differences in grand averaged MRCPs between the initial and final closed-loop BMI therapy sessions. MRCPs were calculated with respect to movement onset times identified from EMG activity of the impaired hand. For this, EMG signals were denoised using Teager-Kaiser energy operator, low-pass filtered (0.5Hz, 4^th^ order Butterworth), standardized, and then compared against a threshold of 0.5 standard deviation to identify intervals of either flexor or extensor contraction (Tenan et al., 2017). Contraction intervals larger than 1 second were retained for further analysis and their time of onset was utilized to segment EEG epochs for calculating MRCPs. This approach ensured that the MRCPs were measured with respect to true movement onset and independent of the classifier’s predictions. To obtain a sufficient number of trials for averaging, we combined EEG epochs from the first 2 and final 2 therapy sessions and then computed their difference. Further, we looked at difference in MRCP peak amplitudes and latency from scalp EEG electrodes located over the motor cortex, specifically, central (C_z_, C_1_- C_4_), fronto-central (FC_z_, FC_1_ – FC_4_) and centro-parietal electrodes (CP_z_, CP_1_ – CP_4_). Further, to account for left hand vs. right hand impairment, the electrode locations were flipped for individuals with right hand impairment. Finally, MRCP latency was defined as time difference starting from 50% of peak amplitude until the time of movement onset (see supplementary materials, Fig. S1) (Müller-Gethmann et al., 2000).

### E Data Analysis and Statistics

The benefit of BMI-enabled exoskeleton therapy was assessed with two objectives, namely improvement in patient engagement (measured as a participant’s ability to reliably operate a BMI) and improvement in motor function (measured via changes in neural activity, clinical scores, and movement kinematics). BMI performance was quantified per session in terms of prediction accuracy, false positives, early detection time, and user feedback. Prediction accuracy was determined based on the fraction of successful trials from total trials, while to calculate false positives, we used catch trials that asked participants to intentionally remain idle during those trials. Our early detection time metric measured how far in advance the BMI could predict movement before a participant tried to move their impaired arm (as seen from EMG activity). The participants’ approval rating of the BMI’s decisions was assessed using a 3-point Likert Scale, with a scoring scale of 3 = Approve, 2 = Not sure, and 1 = Disapprove. To compare offline vs. online BMI performance metrics we used Wilcoxon rank sum test, since the data was non-normal and had unequal sample sizes.

To test for statistical significance of motor recovery based on clinical assessments, one-way mixed effects analysis with repeated measures was used. The assessment intervals were taken as fixed effect with four levels (Baseline, post-treatment, 2-weeks, and 2-months follow-up). Whereas a between-subject intercept was considered as the random effect. Mixed effects models were selected over conventional repeated measures ANOVA, to compensate for the missing follow-up sessions (Wainwright, 2007). Additionally, an in-depth analysis of FMA-UE and ARAT subscales was conducted to assess which of their scoring items improved amongst participants and how long were the improvements retained post-intervention.

Movement quality improvements were evaluated by comparing kinematic data from initial 2 with final 2 therapy sessions. These metrics included Average Speed, Spectral Arc Length (a frequency-domain measure that increases in value as movements become less jerky (Balasubramanian et al., 2015)), and two metrics related to the shape of the velocity profile: Number of Peaks (a higher number of peaks corresponds to jerkier movement), and Time to 1^st^ Peak (which is usually less than the ideal value of 0.5, or 50% of the total movement duration, when a movement has more than one peak). Due to the non-normality of the data, Wilcoxon signed rank tests were used on the paired differences for each movement quality metric.

All data analysis were performed in MATLAB R2018b, with the exception of mixed effects analysis which was carried out in R (R Core Team, 2017) and its ‘lme4’ package (Bates et al., 2015). The statistical significance criteria was set at *p*-values less than 0.05.

## 3. Results

### A BMI Performance across Participants

During the 4-6 weeks long therapy regime, on average, participants completed 132 ± 22 repetitions per session by triggering the exoskeleton’s movement via the BMI. As seen in Fig. 3A top plot, the average BMI’s prediction accuracy was consistently better than random chance (= 50%) across sessions. Inter-subject variability in accuracy reduced with training and during the last 5 therapy sessions, 4 participants achieved greater than 90% accuracy. Overlaid on the plots are BMI performance traces for participants with best (P9) and worst (P7) accuracies across sessions. The remaining plots in this figure demonstrate the BMI’s performance in terms of its ability to avoid false positives, its early detection time, and users’ approval rating. The dotted lines are best fit lines for which the slope was non-zero and statistically significant (*p* < 0.05).

**Fig. 3.**
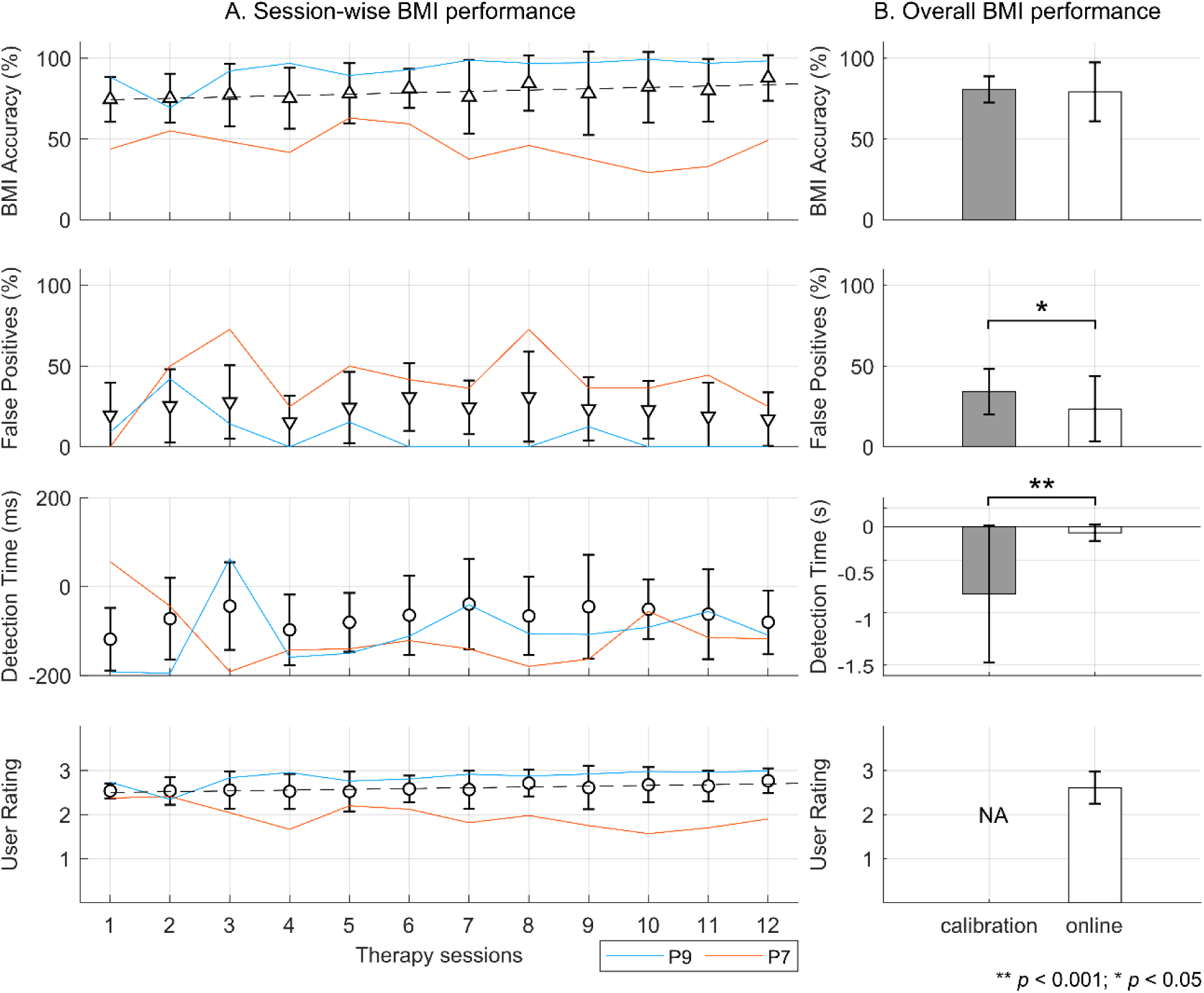
BMI performance in 10 chronic stroke survivors over 12 therapy sessions, averaged by session in sub-plot A and averaged by online testing vs. calibration in sub-plot B. From top to bottom, mean ± s.d. values for BMI’s prediction accuracy, false positives, early detection time, and user approval rating are shown. Results from 2 participants (P9 and P7) with best and worst BMI accuracy are overlaid on the plots. Dotted lines indicate statistically significant trends in accuracy and user rating.

Fig. 3B compares the BMI’s online performance with its offline performance at calibration. Overall the BMI’s accuracy was similar during online and offline testing (79 ± 18% vs. 81 ± 8%, n.s.). The average false positives in the online scenario were significantly smaller than offline (23 ± 20% vs. 34 ± 14%, *p* < 0.05). In offline testing, motor intent could be detected as early as 723 ± 740ms before onset of movement, while in the online case the early detection of intent could be made only 66 ± 86ms in advance (p < 0.001). Finally, the average approval rating was high and consistent across users at 2.6 ± 0.4 points on a 3-point Likert scale.

### B Clinical Outcomes

Fig. 4 shows changes in clinical metrics from baseline evaluated at different time points: post-treatment, 2-weeks, and 2-months follow-ups. The average change in FMAUE and ARAT during the entire course of the study were 3.92 ± 3.73 and 5.35 ± 4.62, respectively. Repeated measures mixed effects model analysis confirmed that there were significant improvements from baseline in FMA-UE (F (23.03, 3) = 5.54, *p* < 0.01) and ARAT (F (23.018, 3) = 6.25, *p* < 0.01). Post-hoc analysis revealed that FMA-UE and ARAT scores after treatment and at follow-ups where significantly better than at baseline. Moreover, as shown in. Table S2 (supplementary materials), overall 8 participants achieved minimal clinically important difference (MCID) after therapy or at follow-ups, based on their FMA-UE and ARAT scores. MCID thresholds for FMA-UE was set as 5.2 points and for ARAT as 5.7 points change from baseline (Lee et al., 2001; Page et al., 2007). No change in JTHFT scores was observed. Marginal improvements in grip and pinch strengths were noted, but these did not reach statistical significance.

**Fig. 4.**
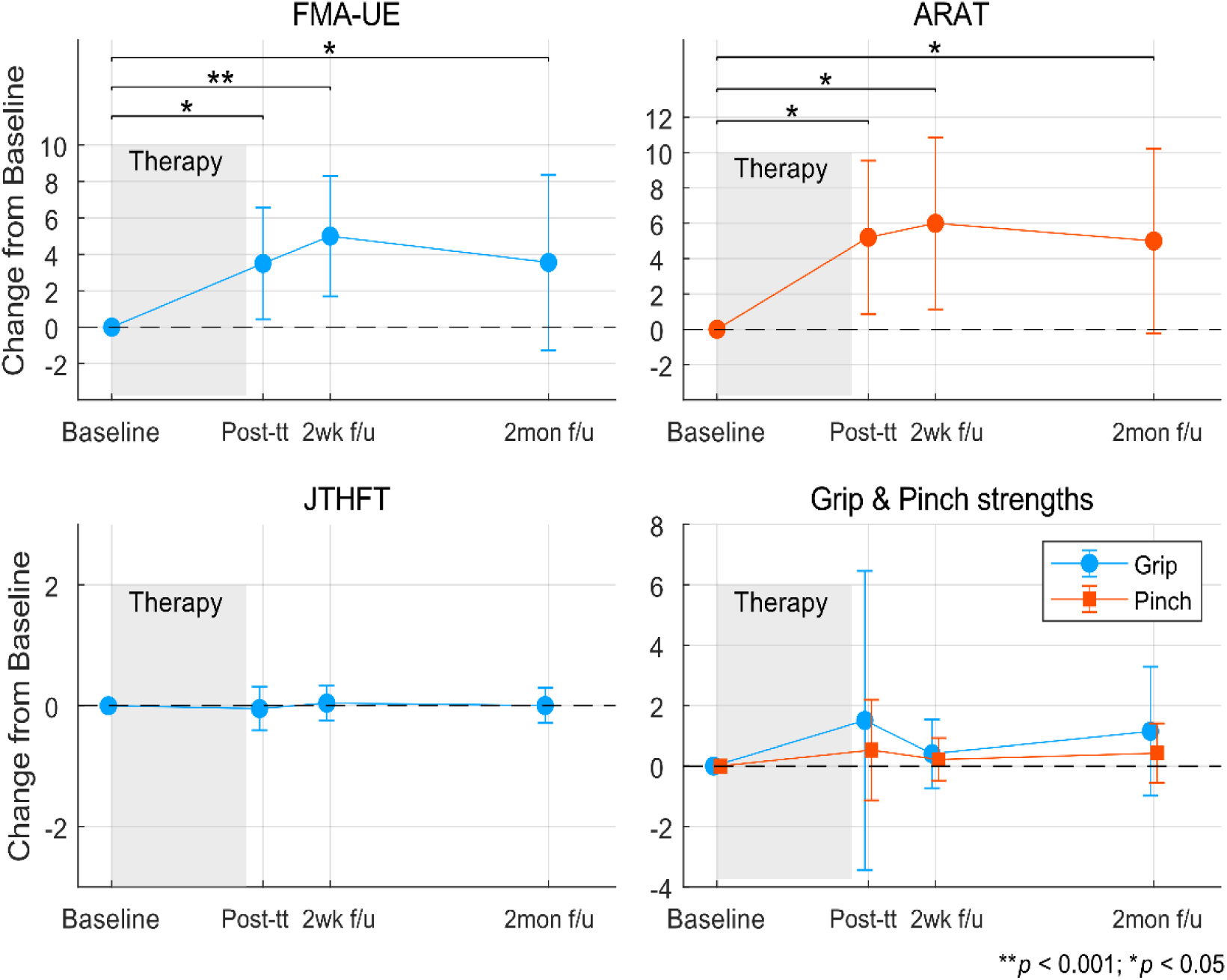
Clinical outcome metrics assessed post-treatment (post-tt) and at 2-week (2wk f/u) and 2-months (2mon f/u) follow-ups relative to baseline. Shaded regions indicate the 4 – 6 weeks long intervention period.

#### i. Changes in FMA-UE and ARAT scores by subscales

In Fig. 5, we breakdown the FMA-UE and ARAT scores into its constituent subscales. For each of the spider charts shown in the figure, the black outer polygon represents maximum score achievable under each subscale. The maximum score in each scoring item is also stated next to each vertex in subplots A & B, as well as in all remaining subplots. The colored polygons represent the 4 different assessment time points, namely baseline, immediately after treatment, 2-weeks and 2-months follow-ups.

**Fig. 5.**
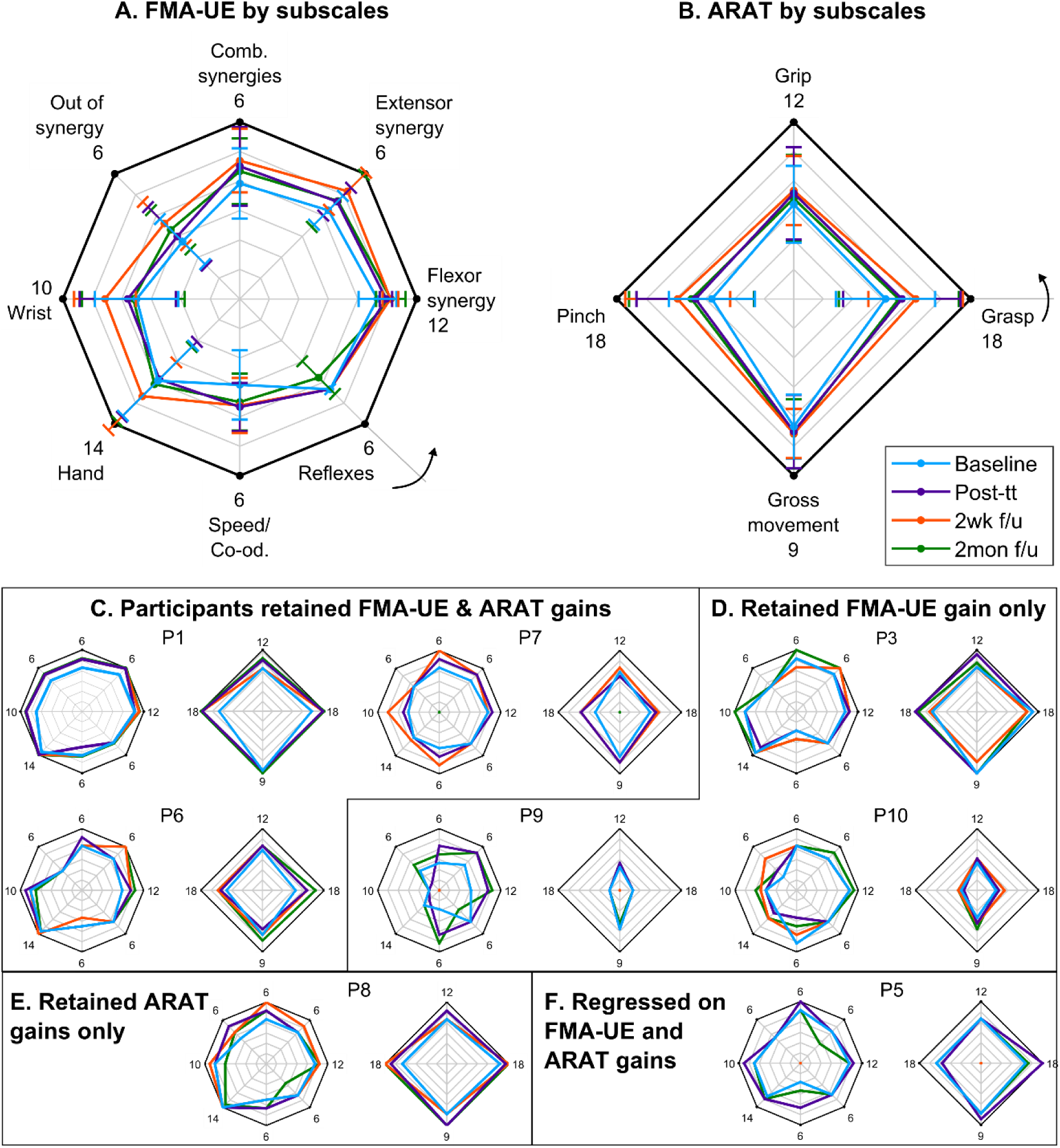
Breakdown of FMA-UE and ARAT scores by subscales, shown by averaging across participants (subplot A & B) and individually (subplots C-F) for participants that achieved minimal clinically important difference. Subplots C-F, further group participants based on their FMA-UE and ARAT outcomes at 2 months follow-up. The arrows in subplots A & B indicate the order of administering the test, starting at the first item and then progressing counter-clockwise.

Fig. 5A & Fig. 5B show the mean ± s.d. scores for FMA-UE and ARAT subscales. On average participants improved in movements involving arm synergies, speed, coordination, wrist and hand components of FMA-UE, as well as grasp and pinch components of ARAT. The improvements were greatest at 2-weeks assessment, but later regressed and at 2-months follow-up the scores were similar to that of post-treatment, albeit better than baseline. Subplots C-F in Fig. 5 track progress of individual participants that were able to achieve MCID during any of the follow-up assessments. For participants that did not attend a follow-up visit (i.e. P5, P7, and P9), their score was assigned zero in the plots and their most recent assessment score were used for further groupings. Specifically, subplot C groups individuals that retained gains in both FMA-UE and ARAT scores at 2-months follow-up (with the exception of P7). Fig. 5D groups individuals that retained gains in FMA-UE, but either regressed or did not improve their ARAT scores. Similarly, Fig. 5E shows a participant who retained his ARAT scores, but regressed on FMA-UE. Finally, subplot F shows a participant that regressed on both FMA-UE and ARAT scales at 2-months follow-up.

### C Behavioral Outcomes

#### i. Motion Kinematics

In Fig. 6, boxplots compare movement quality metrics between the start and end of therapy sessions. Using a single-sided Wilcoxon signed rank test, the median values for Average Speed, Spectral Arc Length, Number of Peaks and Time to 1^st^ Peak were significantly higher at the end of the therapy. Median values for Average Speed increased from 13.6 deg/s to 23 deg/s (*p* < 0.05) and Spectral Arc Length increased from -2.29 to -2.17 (p < 0.05). The median Number of Peaks decreased from 2.11 to 1.68 (*p* < 0.001), which suggests that movements at the end of therapy were less jerky. Also, the median Time to 1^st^ Peak increased from 0.36 to 0.45 (*p* < 0.001), which indicates well-balanced movements (ideal value = 0.5) were achieved post-therapy completion.

**Fig. 6.**
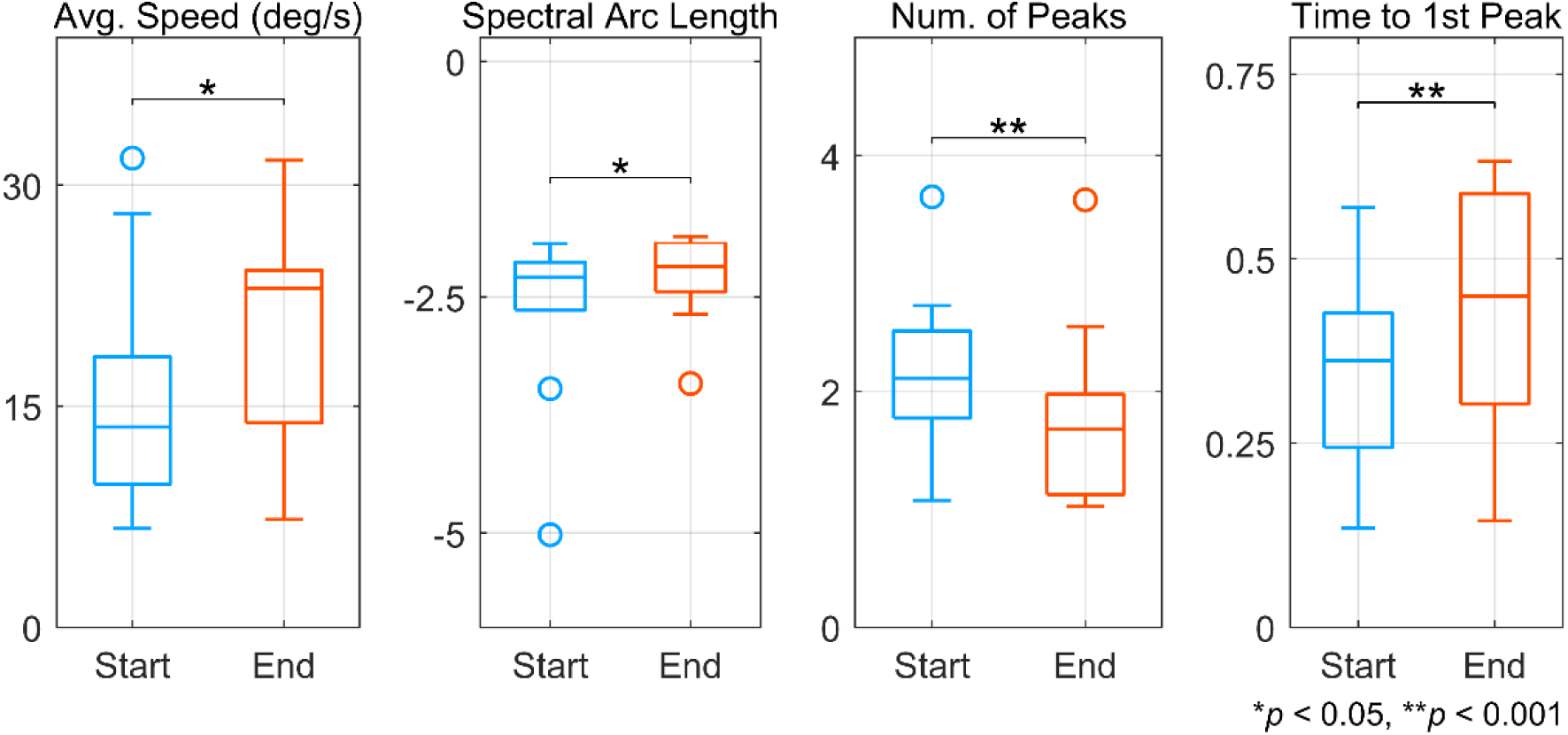
Improvement in movement quality between start and end of therapy. Movement quality was derived from joint angle velocity using various kinematic metrics. For all metrics except Number of Peaks, an increase in value corresponds to improvement.

#### ii. Presence of Global Synkinesis

Bilateral surface EMG analysis revealed that the involuntary co-activation of unimpaired arm when using paretic arm, also known as global synkinesis phenomenon, existed in 2 participants: P4 (baseline FMA-UE = 21, ARAT = 4) and P8 (baseline FMAUE = 49, ARAT = 42). As seen from the normalized bilateral EMG traces in Fig. S2 (supplementary materials), synkinesis was primarily observed during elbow extension, while it was absent during elbow flexion. Moreover, no change was observed in the extent of co-activation of the unimpaired arm between the start and end of therapy.

### D Correlation of MRCP Amplitude and Latency with Clinical Outcomes

We correlated changes in FMA-UE and ARAT scores post-treatment with differences in MRCP signals, corresponding to initial and final therapy sessions. As seen in Fig. 7 top row, MRCP amplitude from the contralateral EEG electrodes highly correlated with functional assessment scores. Specifically, change in average MRCP amplitude for contralateral central electrode (i.e. C_1_ or C_2_ depending on impaired side and abbreviated as C_1/2_) significantly correlated with ARAT scores (ρ = 0.72, *p* < 0.05). Likewise, correlation between MRCP amplitude from contralateral fronto-central electrode (i.e. FC_1_ or FC_2_ depending on impaired side and abbreviated as FC_1/2_) and FMA-UE scores, was tending towards significance (ρ = 0.63, *p* = 0.051). No significant correlation between MRCP latencies and clinical outcomes was observed. The bottom row in Fig. 7A & *Fig*. 7B, plots the averaged MRCP signals from the initial and final therapy sessions for all participants, corresponding to central and fronto-central EEG electrodes.

**Fig. 7.**
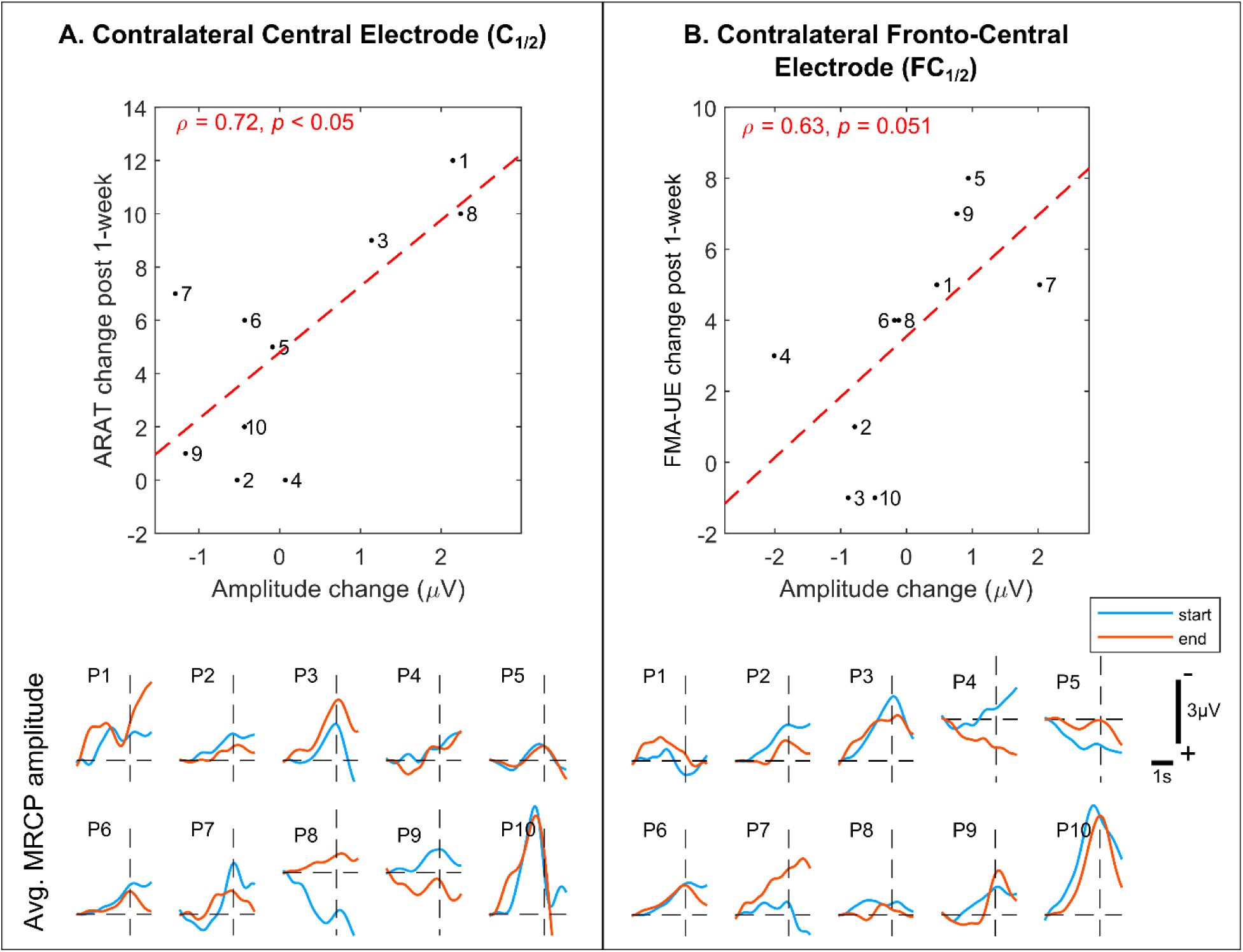
Correlation (*ρ*) between MRCP amplitude and functional assessment scales. Top row compares MRCP amplitudes from central and fronto-central EEG electrodes with clinical outcomes. In these figures, numbers represent participant I.Ds. Bottom row shows MRCPs recorded from all the participants at start and end of therapy. Note, MRCPs are aligned with respect to movement onset (t = 0s).

## 4. Discussion

Cortical reorganization and motor recovery following stroke are contingent on ensuring active user engagement and participation during rehabilitation, to promote activity-dependent neuroplasticity (Venkatakrishnan et al., 2014). Towards this extent, BMI-based neurorehabilitation therapies have performed arguably better at engaging the user and achieving better functional outcomes than any other contemporary rehabilitation therapies (e. g. CIMT, robot-assisted or neuromuscular stimulation alone, etc.) (Cervera et al., 2018). In the same light, this study confirmed that BMI-enabled robot-assisted upper-limb therapy resulted in improved motor function for a majority of the participants with chronic stroke, as determined from post-treatment, 2-weeks, and 2-months assessments.

Specifically, functional metrics that are typically associated with arm/hand movements and co-ordination, i.e. FMA-UE and ARAT, improved as a result of therapy (7 participants with moderate-mild impairment and 1 with severe-impairment showed some level of motor recovery by the end of the intervention). Whereas, metrics associated with hand strengthening and speed, such as JTHFT, grip and pinch strengths remained stable. Since the BMI-enabled MAHI Exo-II exoskeleton was primarily targeting elbow training, this result is expected. However, as seen in Fig. 5A & Fig. 5B, the effects of elbow training generalized to positive improvements in wrist and hand subscales of FMA-UE and pinch and grasp subscales of ARAT outcomes. Moreover, no adverse events directly related to the intervention were reported, although one participant (P5) experienced unexplained tiredness, forgetfulness, and excessive decline in motor performance, 2 months after therapy (see Fig. 5F).

While clinical outcomes are indisputable evidence of motor relearning, often these are imperceptive to cortical changes at sub-clinical levels. Hence, to determine the efficacy of any neurorehabilitation therapy, it is important to identify neural correlates or biomarkers that can explain and even predict post-treatment clinical outcomes. Indeed, previous studies have identified neural correlates based on the BOLD response (Ramos-murguialday et al., 2013), white matter tract anisotropy (Song et al., 2015), and brain symmetry index (Ang et al., 2014). Our analysis of MRCPs from start to end of therapy showed that participants who improved in motor function were characterized by changes in the MRCP amplitudes from the contralateral EEG electrodes that were highly and positively correlated with functional assessment scores. More specifically, MRCP amplitudes from the primary motor cortex and supplementary motor area (Brodmann Areas 4 & 6) contralateral to the impaired arm, correlated with ARAT (ρ = 0.72, *p* < 0.05) and FMA-UE (ρ = 0.63, *p* = 0.051) scores, respectively (Koessler et al., 2009). However, no significant correlation with MRCP latency was observed. Since MRCP amplitude is believed to encode information about computational effort and attention (Cui & Mackinnon, 2009), increase in MRCP amplitude suggests higher activation of the ipsilesional hemisphere. Thus, MRCPs can serve as a viable neural correlate for predicting clinical outcomes of any neurorehabilitation therapy.

Interestingly, even though our participants performed a small number of physical movements per session (132±22), their functional and kinematic outcomes were comparable to high-intensity robot-only therapies (Klamroth-Marganska et al., 2014; Lo et al., 2010). This was likely facilitated by the BMI’s consistent decoding accuracy (avg. = 79 ± 18%), low false positives (23 ± 20%) and early detection latency (−66 ± 86ms). This in turn allowed the exoskeleton to seamlessly respond to the participant’s volitional movement intent and provide causal afferent sensory feedback, thereby promoting cortical plasticity.

Our study did have a few shortcomings. Firstly, the absence of a control group prevented us from understanding the individual benefits of BMI and robotic therapy alone. However, we ensured that the participants enrolled had a stable baseline and any improvements can be attributed to the combined effect of BMI plus robotic therapy. Secondly, our sample size was small (n = 10), which prevents us from generalizing the outcomes to a larger sample. This was in part to our narrow inclusion criteria, which excluded about 75% of the participants that were screened. Lastly, the BMI control was limited to one-dimensional (Go vs. No-go), which might not have been engaging enough for some of the participants (e.g. P7). For future participants, it will be our priority to achieve multi-dimensional BMI control and combine it with virtual or augmented reality, to provide an immersive learning environment. Nonetheless, our study found compelling evidence and an MRCP-based neural correlate for clinical efficacy of BMI-enabled robot-assisted rehabilitation.

## Data Availability

Data is available from the corresponding author upon reasonable requests.

## Notes

### Competing Interest Statement

The authors have declared no competing interest.

### Clinical Trial

NCT01948739

### Funding Statement

This work was supported by NIH National Robotics Initiative Grant R01NS081854 and a grant from Mission Connect, a project of TIRR Foundation.

